# Use of wastewater metrics to track COVID-19 in the U.S.: a national time-series analysis over the first three quarters of 2022

**DOI:** 10.1101/2023.02.06.23285542

**Authors:** Meri Varkila, Maria Montez-Rath, Joshua Salomon, Xue Yu, Geoffrey Block, Douglas K. Owens, Glenn M Chertow, Julie Parsonnet, Shuchi Anand

## Abstract

**Background:** Widespread use of at-home COVID-19 tests hampers determination of community COVID-19 incidence. Using nationwide data available through the US National Wastewater Surveillance System, we examined the performance of two wastewater metrics in predicting high case and hospitalizations rates both before and after widespread use of at-home tests.

**Methods:** We performed area under the receiver operating characteristic (ROC) curve analysis (AUC) for two wastewater metrics—viral concentration relative to the peak of January 2022 (“wastewater percentile”) and 15-day percent change in SARS-CoV-2 (“percent change”). Dichotomized reported cases (≥ 200 or <200 cases per 100,000) and new hospitalizations (≥ 10 or <10 per 100,000) were our dependent variables, stratified by calendar quarter. Using logistic regression, we assessed the performance of combining wastewater metrics.

**Results:** Among 268 counties across 22 states, wastewater percentile detected high reported case and hospitalizations rates in the first quarter of 2022 (AUC 0.95 and 0.86 respectively) whereas the percent change did not (AUC 0.54 and 0.49 respectively). A wastewater percentile of 51% maximized sensitivity (0.93) and specificity (0.82) for detecting high case rates. A model inclusive of both metrics performed no better than using wastewater percentile alone. The predictive capability of wastewater percentile declined over time (AUC 0.84 and 0.72 for cases for second and third quarters of 2022).

**Conclusion:** Nationwide, county wastewater levels above 51% relative to the historic peak predicted high COVID rates and hospitalization in the first quarter of 2022, but performed less well in subsequent quarters. Decline over time in predictive performance of this metric likely reflects underreporting of cases, reduced testing, and possibly lower virulence of infection due to vaccines and treatments.

## Introduction

Rapid determination of COVID-19 incidence within communities can guide screening at hospitals, residential facilities, schools, or communal gatherings, mobilize treatment supplies, and preserve hospital capacity. Public health agencies including the US Centers for Disease Control and Prevention (CDC) rely chiefly on rates of reported new COVID-19 cases and/or hospitalizations to estimate county ‘level’ of COVID-19^1^. As home testing becomes widespread, however, case counts are likely to substantially underestimate incidence, to a degree depending on at-home test availability, acceptance, and cost, as well as the severity of disease seen with circulating strains of SARS-CoV-2^2^. Similarly, since the introductions of vaccines and medications that reduce COVID-19 severity^3^, tracking hospitalization rates may produce unreliable estimates of disease incidence. The lack of accurate data regarding community burden leaves high-risk patients at particular risk.

Wastewater surveillance offers a potential solution to the problem of accurate SARS-CoV-2 surveillance—a solution agnostic to symptomatic, diagnosed, or reported disease. High-resolution sequencing of wastewater can also identify emerging variants of concern^4^ and estimate effective reproductive number^5^, a key predictor of future transmission. For these reasons, many jurisdictions are investing in expanding wastewater surveillance, with over 70 countries and more than 3500 sites reporting data to a central dashboard^6^. Yet adoption of wastewater surveillance to inform public policy has not yet become widespread, in part due to challenges interpreting results with shifting detection methods, strains, populations served, and wastewater dynamics^7^. To date, available data evaluating wastewater metrics against cases or hospitalizations in the US are geographically limited, evaluating a single sewershed^8,9^ or a few sewersheds grouped regionally^10^.

The CDC’s National Wastewater Surveillance System (NWSS) collates data from a majority of currently operating wastewater testing sites in the US^11^. The NWSS normalizes wastewater samples to wastewater flow and population size served and relies on viral gene copies per individual in the sewershed as the foundational data unit. This normalization addresses concerns about changes due to weather and differences in sewershed size. Despite normalization and generation of aggregate measures (e.g., percent change in normalized virus concentration in the last 15 days), no interpretation algorithm is provided to inform screening policy. Indeed, the NWSS specifically recommends that “point estimates of community infection based on wastewater measurements …not be used” to shape policy, largely because the amount of virus shed by individuals with infection into the sewage system has not been well characterized^12^. Yet, with decreased institutional testing and lower disease virulence for a majority of the immunocompetent disease population, wastewater may be the best (and possibly only) way to understand dynamics of circulating SARS-CoV-2 virus in communities.

Using data from the NWSS, we sought to evaluate how well national data on wastewater SARS-CoV-2 measures paralleled reported new COVID-19 cases and hospitalizations over time in the US. We hypothesized that the correlation between wastewater and COVID-19 disease metrics would be relatively strong prior to widespread home COVID-19 testing and would weaken over time; i.e., that there would be attenuation of the association between viral transmission and formally reported new case and hospitalization rates. Our overall goal was to determine whether wastewater surveillance is preferable to currently employed disease metrics to inform public policy around SARS-CoV-2 screening and resource allocation.

## Methods

### Data sources

#### Wastewater metrics

We obtained publicly available data from the NWSS spanning the Omicron variant dominant period of January-September 2022^11^. The NWSS reports data on wastewater from public health department-monitored sewersheds serving at least 3000 people. Excluded are sewersheds missing population estimates, or that represent single institutions (e.g., a university). The sewersheds quantify SARS-CoV2 in unconcentrated wastewater or sludge using either reverse transcription quantitative polymerase chain reaction (RT-qPCR; 672 sites) or reverse transcription digital polymerase chain reaction (RT-dPCR); 545sites); irrespective of sample type and methodology, sites report virus concentrations per volume^12^. Publicly reported data include sewershed location identifiers and population served. In an effort to facilitate comparisons, aggregated metrics include facility SARS-CoV-2 percentile (i.e., an ordered rank of the current virus concentration relative to historic peak and nadir at that facility; “wastewater percentile”), percent change in normalized virus concentration in the prior 15 days (“percent change”), and percent of wastewater samples with detectable virus in the prior 15 days. Our analysis focused on the first two measures.

Because the wastewater percentile variable compares current to historical viral concentrations, and since we aimed to compare data across sites, we restricted our analysis to sewersheds with available data in January 2022, the period of the highest community transmission rates of COVID-19 in the US^13^ (Figure S1). January 2022, the peak of original (BA 1.1) Omicron strain, was also the peak period for laboratory and point-of-care testing; positive tests would likely have been reported to health authorities. Home tests were shipped free to households in the US starting at the end of January^14^.

Counties had to have submitted at least one week of data in January 2022 and have data available for >50% of subsequent weeks in order to be further included in the analysis. We also restricted data to samples obtained from a treatment plant itself rather than from pre-treatment plant wastewater. In counties with more than one sewershed reporting to NWSS, we aggregated data from each sewershed to county level by creating a weighted average using each sewershed’s population served. Thus, the sewershed serving the largest population contributed the largest weight to the averaged county estimate.

#### Case and hospitalization rates

In the primary analysis, we employed two CDC community level indicators as our dependent variables: reported new COVID-19 cases per 100,000 and new inpatient admissions per 100,000^15^. We used publicly available time series data on aggregated counts of COVID-19 cases from state and local health departments, and hospital admissions from U.S. Department of Health and Human Services Unified Hospital Data Surveillance System^16,17^. Consistent with CDC reporting practices, we computed aggregate counts of COVID-19 cases and hospitalizations per 100 000 population from the past seven days at the midpoint of each week. When comparing wastewater metrics to hospitalizations, we lagged new inpatient admissions by two weeks. We defined high COVID-19 community level using CDC-defined thresholds:

- Reported case rate equal to or greater than 200 new COVID-19 cases per 100 000 population
- Reported hospitalization rate equal to or greater than 10 new inpatient admissions per 100 000 population

### Statistical analysis

We grouped data by calendar quarters (January-March, April-June, and July-September). We obtained county population data from the 2021 US Census^18^. To visually evaluate the correlation between our two wastewater metrics and clinical case metrics, we graphed these for 2022 for the most populous county from each US Census region. We additionally graphed the absolute wastewater concentrations within the county to visualize its association with the county-level wastewater metrics.

We then computed the sensitivity, specificity, and Area Under the Receiver Operating Characteristic (AUROC) by time period of wastewater metrics in identifying CDC thresholds for high COVID-19 case and hospitalization rates. We treated the wastewater metrics as the ‘test’ of interest and the thresholds of cases and hospitalizations as comparative indicators of high COVID-19 community level. To determine whether combining both wastewater metrics could be used to predict high COVID-19 community levels, we used logistic regression accounting for wastewater percentile, percent change, and the interaction of the two. In a sensitivity analysis, we evaluated wastewater percentile predictive performance in small versus large counties; large counties are defined as population equal to or greater than 500,000 by US Census Bureau. We also conducted sensitivity analysis using current cases and hospitalization rates as predictors, and rates of both lagged by two weeks above CDC thresholds as our dependent variables.

Each county contributed data to the analysis for weeks during which wastewater data was available. Because not all counties consistently reported values, the number of counties included in the analysis varies per week. We assessed the relationship between the wastewater and clinical case metrics for each week stratified by calendar quarter. We computed bootstrapped confidence intervals for sensitivity at given specificity points using the *ci.se* function in R (version 4.2.2, R Foundation for Statistical Computing).

We used R statistical packages epiR and pROC to perform the analyses.

## Results

Among 730 wastewater treatment counties that submitted the analyzed metrics to NWSS during our study period, 268 counties across 22 states met our inclusion criteria (Figure 1, Figure S2). Median population of counties included in the analysis was 95,938 (25^th^, 75^th^ percentile 44,697, 294,772) residents (Table 1). Comparatively the overall US county population median is 25,752 (25^th,^ 75^th^ percentile 10,818, 67,899). Consistent with national data, reported new case and hospitalization incidence rates were high in the first and third quarters, and lower in the second. Also consistent with national data, in our sampled counties, new case incidence was at its highest since the start of the pandemic in January 2022 (Figure S2).

**Table 1.** Sampled county population and sewershed data, and reported case and hospitalization rates by calendar quarters of 2022.

|  | **Counties**<br>**(n=268<sup>a</sup>)** |  |
| --- | --- | --- |
| Total County Population, median (IQR) | 95,938 (44,697, 294,772) |  |
| Population served by sampled sewersheds, median (IQR) | 49,831 (14292, 221250) |  |
| Sampled sewersheds per county, median (IQR) | 1 (1, 2) |  |
| Weeks with available data<sup>b</sup>, median (IQR) | 39 (38, 39) |  |
| *Calendar quarters of 2022* | **Reported new cases<sup>c</sup>,**<br>**median (IQR)** | **Reported new hospitalizations<sup>c</sup>,**<br>**median (IQR)** |
| Jan-Mar | 246 (73, 928) | 12 (4, 28) |
| Apr-Jun | 132 (60, 211) | 5 (2, 9) |
| Jul-Sept | 180 (129, 238) | 10 (5,16) |
| Data are in median (25<sup>th</sup>, 75<sup>th</sup> percentile) per county. |  |  |
| <sup>a</sup> Number of counties for quarter 3 (July-Sept ) was smaller at 263 |  |  |
| <sup>b</sup> The total study period from January 1<sup>st</sup> 2022 until September 30<sup>th</sup> 2022 spanned 39 calendar weeks |  |  |
| <sup>c</sup> Number of reported new cases and reported new hospital admissions per 100,000 population |  |  |

**Figure 1.**
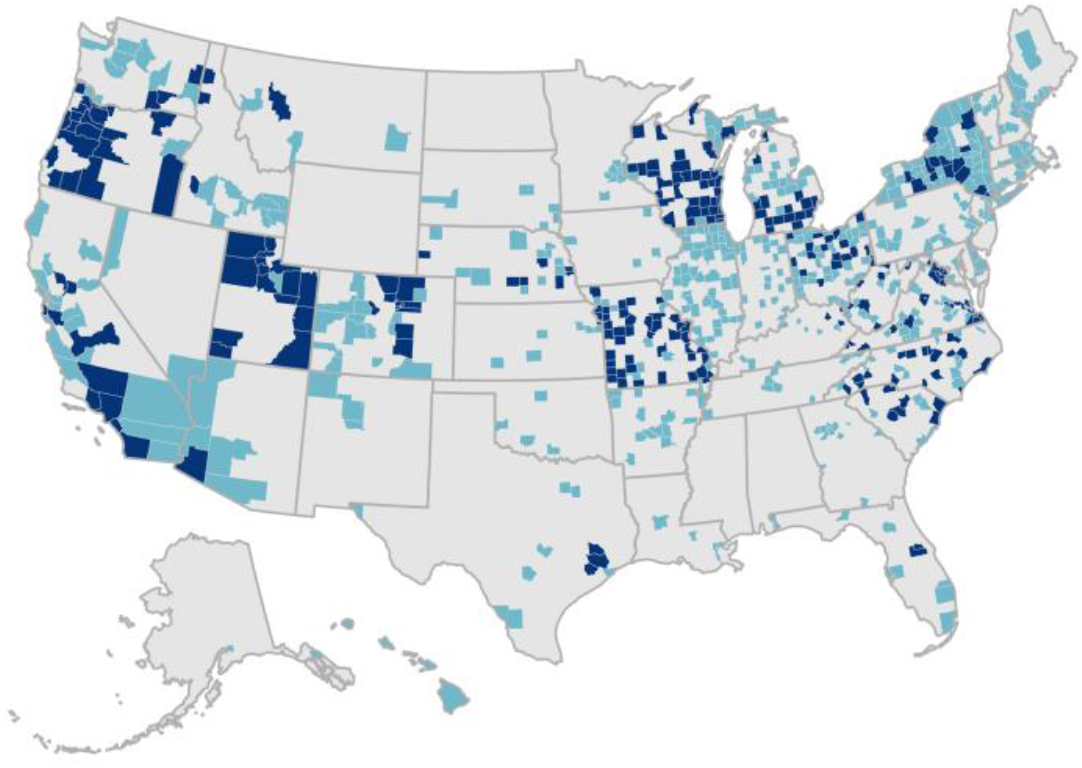
U.S. counties submitting wastewater surveillance data to the NWSS between January 1 2022 and September 30 2022. Regions mapped in dark blue show counties included in the analysis (n=268). Regions mapped in light blue show counties that submitted the analyzed metrics to NWSS, but did not meet inclusion criteria and were excluded from analysis (n=462).

Plots of the available data for 2022 from the most populous counties in each US Census region demonstrate a direct correlation between facility percentile and absolute SARS-CoV-2 concentrations (Figure 2). The 15-day percent change variable fluctuated widely. In the first quarter of 2022, facility wastewater percentile correlated closely with cases and hospitalizations. In contrast, the correlation was less evident in the third quarter when reported case and hospitalization rates were low despite high levels of SARS-CoV2 in wastewater as indicated by facility percentile.

**Figure 2.**
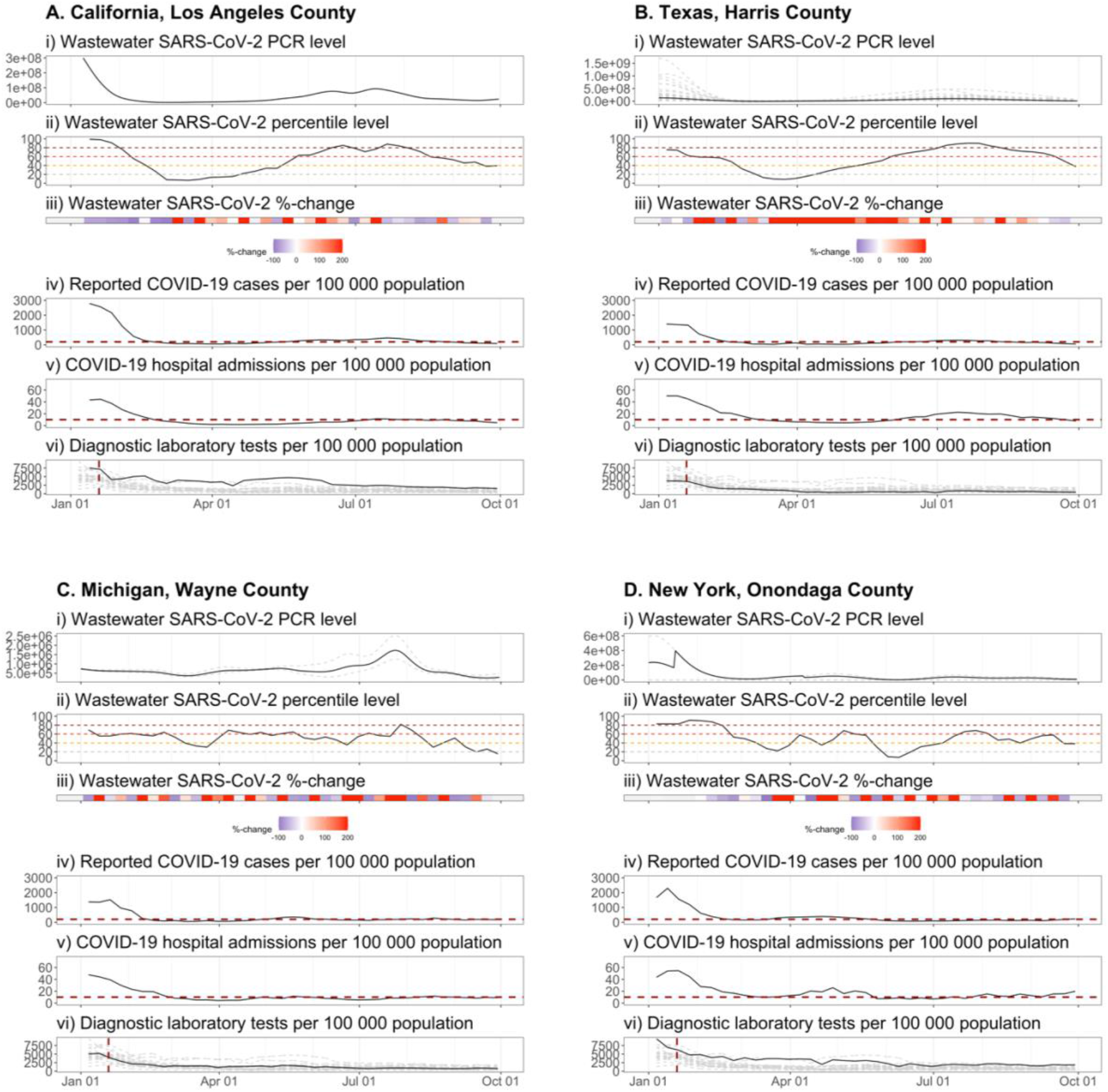
Time history of wastewater surveillance data and clinical case metrics from the most populous counties in each US Census region between January 2022 and September 2022. Data are shown for the most populous counties in U.S. Census regions (A) West: Los Angeles County, California, (B) South: Harris County, Texas, (C) Midwest: Wayne County, Michigan, and (D) Northeast: Onondaga County, New York. Panel (i) shows smoothed spline-fit PCR concentrations of SARS-CoV-2 for each sampling location as reported by the CDC NWSS. When multiple sewersheds were sampled within a county, dashed grey lines in panel (i) represent individual sewersheds. The solid black lines in panels (i), (ii) wastewater SARS-CoV-2 percentile level, and (iii) wastewater SARS-CoV-2 15-day percent change show weighted mean values using each sewershed’s population served. Horizontal dashed lines in panels (iv) and (v) show thresholds for high COVID-19 community level (reported COVID-19 case rate equal to or greater than 200 per 100,000 population and reported hospitalization rate equal to or greater than 10 new inpatient admissions per 100,000 population, respectively). Panel (vi) shows state-level data (solid black lines show reported tests from the state of California (A), Texas (B), Michigan (C), and New York (D); dashed grey lines show estimates for all other U.S. states).

In AUROC analyses incorporating data from all counties, facility wastewater percentile demonstrated excellent predictive power for high reported case (>200 per 100,000) and hospitalization rates (>10 per 100,000 lagged by two weeks) in the first quarter of 2022 (Figure 3). Percentile of 51% and 54% had maximized sensitivity (0.93, 0.80) and specificity (0.82, 0.78) for detecting cases exceeding 200 per 100,000 and hospitalizations exceeding 10 per 100,000, respectively in the first quarter of 2022. Performance was similar in large and small counties (AUC for first quarter 0.95 and 0.95 for cases and AUC 0.85 and 0.94 for hospitalizations in small and large counties respectively, Figures S3-4). In the overall analysis, AUC declined over the next two quarters for predicting both of these clinical thresholds.

**Figure 3.**
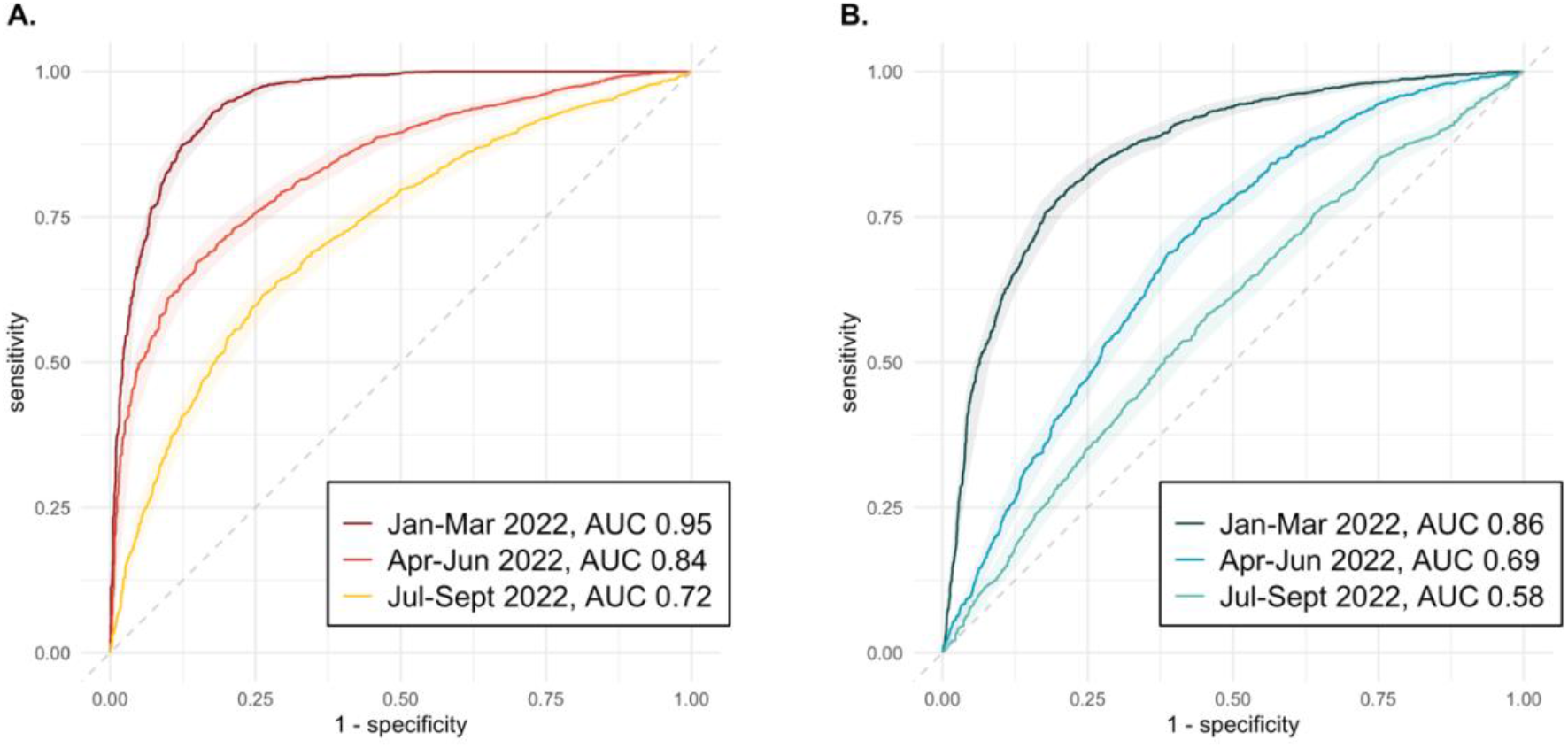
Performance of wastewater percentile in reference to clinical case metrics stratified by calendar quartile of 2022. AUC of wastewater percentile in reference to (A) Current reported COVID-19 cases (≥200 per 100,000 population), (B) New hospital admissions in two weeks (≥10 per 100,000 population).

The percent change metric performed poorly with AUC ranging from 0.51 to 0.57 for reported new cases, and 0.50 to 0.55 for hospitalizations across the three quarters (Figure S5). Combining wastewater facility percentile, percent change, and the interaction of the two in a logistic regression analysis yielded similar estimates of predictive performance as using the percentile metric alone (Figure S6). AUROC analyses examining the performance of clinical metrics as predictors of future clinical outcomes (i.e., do current case and hospitalization rates predict case and hospital admission rates lagged by two weeks) indicated strong predictive power (Figures S7-S8) for both clinical metrics in the first quarter of 2022. As with wastewater percentile, however, predictive performance declined over the next two quarters.

## Discussion

In this first national-level evaluation of CDC-generated wastewater metrics from sewersheds located throughout the nation, we observed a strong direct correlation between a county’s SARS-CoV-2 wastewater concentration relative to its maximal observed, and COVID-19 cases and hospitalizations for US counties during the first quarter of the 2022. When little home testing was being conducted, wastewater percentile in all counties tracked quite closely with new cases per 100,000. However, the correlation of the wastewater percentile with COVID-19 reported cases weakened over the next two quarters, indicating an increasing dissociation between community viral prevalence and reports of infection to health departments. There was also increasing dissociation between wastewater and new hospitalization rates, perhaps indicative of lower rates of COVID-related hospitalization with heightened population immunity due to prior infection and vaccination, potentially lower virulence of evolving strains, and/or reduction in routine admissions testing in hospitals.

In the first quarter of 2022, we evaluate wastewater metrics against case metrics when many wastewater facilities were contributing data nationwide, and when at-home testing, while increasing, was still not ubiquitous^19^. No nationally representative data exist on the relative use of home tests versus laboratory or point-of-care tests in the US, but from a large internet survey of more than 450,000 US adults, among persons with symptoms, there was a 4-fold increase in report of home COVID-19 tests, from 5% in the Delta-dominant period in Fall 2021 to 20% in the Omicron dominant period in Winter 2022. Results of home testing are rarely reported. In a recent analysis of self-testing data from October 2021-May 2022, Ritchey et al. reported results from only 3% of the nearly 400 million at-home tests produced by four US manufacturers were voluntarily reported to health authorities^14^. Since we tracked the same facilities in the same counties over the subsequent two quarters, we postulate that it is the increasing use of at-home testing, rather than any changes in the relation between SARS-CoV-2 incidence and shedding into the wastewater system, that led to our observed decline in the performance of wastewater percentile in ‘detecting’ new cases.

In our analysis, the benchmark to January 2022 is critical for interpretation, since the wastewater percentile metric places all newer data relative to the highest community prevalence of COVID19 seen in the county, with the vast majority of counties experiencing their highest or among their highest prevalence in January 2022. As more wastewater facilities come online, benchmarking remains a critical unresolved question. There are several ways to address lack of historical data including imputation models, and adopting references relative to data from neighboring established facilities. Other potential improvements, such as selecting sewersheds that are better representative of counties sampled, could increase the yield of a sentinel surveillance system.

Since Fall 2020 the CDC has invested over $100 million to support wastewater surveillance infrastructure in the US, with the largest share of the investment occurring in August 2022. Although the utility of wastewater surveillance extends beyond COVID-19, national level data are collated and reported only for COVID-19. The NWSS makes significant efforts to convert absolute viral concentration data into comparable measures across and within sites. As of September 2022, 1213 facilities representing 741 counties and 50 states are submitting data, and are requested to do so weekly. In our analysis, > 99% of analyzed facilities had more than half of weeks covered from the past three quarters. Thus, timely nationwide data are available for an increasing number of US residents. Although a few counties publish and publicize their results^20^ to raise public awareness, inform mask wearing, and promote social distancing, we lack a national strategy for the use of wastewater surveillance. If we presume the first quarter correlation we observed between wastewater percentile and new cases likely holds steady, then wastewater percentile of 51% percent of maximum as generated by the NWSS can provide accurate estimates of high disease burden, regardless of reported case counts.

As a counterpoint, as COVID-19 infection evolves clinically for the largest share of the population, in whom there is a potentially lower risk for hospitalization and death with vaccination and the Omicron subvariants, some could argue that investments in capturing ‘true’ prevalence of circulating disease is unnecessary. This is a reasonable trade-off to consider, but needs to be contextualized with two important points. First, medically vulnerable populations such organ transplant recipients^21^, persons on chemotherapy^22^, and persons receiving dialysis^23^ are suboptimally protected by vaccinations and remain at high risk for adverse health outcomes from COVID-19 infection. Thus, awareness of true disease prevalence could promote additional protective measures tailored to these populations and enable earlier treatment. For example, during periods of high disease circulation, universal asymptomatic testing could be offered in long term care facilities, with nirmatrelvir/ritonavir treatment provided early to patients testing positive. Second, even among the general population COVID-19 infection or reinfection has been shown to be associated with adverse health events, including long COVID symptoms and hospitalizations. Moreover, there remain risks of waning immunity and worsening variants.

Our analysis is limited by the need to rely on a subset of facilities with sufficient data to not only track back to a ‘true’ community peak, but also to allow a relatively stable percentile value assigned to an absolute viral concentration over time. Newer facilities may experience significant fluctuations in the relation between absolute viral concentration and assigned percentile, unless they benchmark to a neighboring county reference and/or use imputed historical data. To facilitate potential public health adoption, we also only evaluated metrics available within NWSS, rather than generate *de novo* metrics using raw or normalized wastewater data.

In summary, in this first analysis of wastewater metrics for SARS-CoV-2 incorporating data from the breadth of public health-monitored sewersheds in the US, we find that counties conducting wastewater surveillance and reporting data to the CDC NWSS in the US could use an aggregated measure of the percent of maximum wastewater SARS-CoV-2 concentration to estimate county-level prevalence of COVID-19. Counties with a longer historical data record, tracking back to at least January 2022, will generally provide the most reliable estimates. We demonstrate that wastewater surveillance can be operationalized to fulfill the relevant public policy goals of public awareness of true SARS-CoV-2 incidence and implementation of additional actions specifically designed to protect medically vulnerable populations.

## Data Availability

We used publicly available data for this analysis. The dataset we constructed for this analysis is available upon request to Dr Varkila.

https://github.com/nytimes/covid-19-data

https://healthdata.gov/Hospital/COVID-19-Reported-Patient-Impact-and-Hospital-Capa/anag-cw7u

https://data.cdc.gov/Public-Health-Surveillance/NWSS-Public-SARS-CoV-2-Wastewater-Metric-Data/2ew6-ywp6

## Acknowledgments

We would like to thank Dr. Alexandra Boehm (Stanford University) and Dr. Alexander Yu (California Department of Public Health), and Dr. Jason Andrews (Stanford University) for their assistance with interpretation and analysis of time series wastewater data.

