## Supplemental appendix for "Use of wastewater metrics to track COVID-19 in the U.S.: a national time-series analysis over the first three quarters of 2022"

**Supplementary Table 1** Sampled county population and sewershed data, and case and hospitalization rates by quarters of 2022

**Supplementary Fig. 1** Diagnostic testing and reported new COVID-19 cases in the U.S between March 1 2020 and September 30 2022

**Supplementary Fig. 2** Selection of counties included in analysis

**Supplementary Fig. 3** Performance of wastewater percentile in reference to clinical case metrics in small U.S. counties stratified by calendar quartile of 2022

**Supplementary Fig. 4** Performance of wastewater percentile in reference to clinical case metrics in large U.S. counties stratified by calendar quartile of 2022

**Supplementary Fig. 5** Performance of 15-day wastewater percent change in reference to clinical case metrics stratified by calendar quartile of 2022

**Supplementary Fig. 6** Performance of combined wastewater metrics in reference to clinical case metrics stratified by calendar quartile of 2022

**Supplementary Fig. 7** Performance of current reported COVID-19 case rates in reference to clinical case metrics stratified by calendar quartile of 2022

**Supplementary Fig. 8** Performance of current COVID-19 hospital admission rate in reference to clinical case metrics stratified by calendar quartile of 2022

### Supplementary Table 1. Sampled county population and sewershed data, and case and hospitalization rates by quarters of 2022

| | Jan-Mar 2022 (n=268) | | | | | Apr-Jun 2022 (n=268) | | | | | Jul-Sep 2022 (n=263) | | | | |
|---|---|---|---|---|---|---|---|---|---|---|---|---|---|---|---|
| | Median | Min | Max | 25th | 75th | Median | Min | Max | 25th | 75th | Median | Min | Max | 25th | 75th |
| Weeks with available data, number | 13 | 1 | 13 | 12 | 13 | 13 | 4 | 13 | 13 | 13 | 13 | 8 | 13 | 13 | 13 |
| Wastewater sites per county, number | 1 | 1 | 33 | 1 | 2 | 1 | 1 | 33 | 1 | 2 | 1 | 1 | 33 | 1 | 2 |
| Total county population, | 95938 | 8376 | 9829544 | 44697 | 294772 | 95938 | 8376 | 9829544 | 44697 | 294772 | 96017 | 8376 | 9829544 | 45242 | 299647 |
| County population served by sampled sewersheds | 49831 | 3076 | 3500000 | 14292 | 221250 | 49831 | 3076 | 3500000 | 14292 | 221250 | 49662 | 3076 | 3500000 | 14410 | 198804 |
| Wastewater percentile level, % | 46.6 | 0 | 100 | 17.0 | 83.5 | 44.4 | 0 | 95.8 | 25.0 | 62.2 | 60.5 | 1.50 | 100.0 | 47.7 | 72.2 |
| 15-day percent change variable, % | -42 | -100 | 2147483647 | -80.0 | 51 | 30 | -100 | 2147483647 | -39 | 226 | -8 | -100 | 2147483647 | -60 | 91 |
| New cases per 100,000 population | 245.8 | 3.0 | 4728.6 | 73.3 | 928.1 | 132.1 | 0 | 826.3 | 59.9 | 210.7 | 180.0 | 24.4 | 994.5 | 128.5 | 238.1 |
| New hospitalizations per 100,000 population | 11.5 | 0 | 291.7 | 4.2 | 27.7 | 4.8 | 0 | 70.9 | 2.0 | 9.3 | 9.6 | 0 | 124.2 | 5.4 | 15.5 |

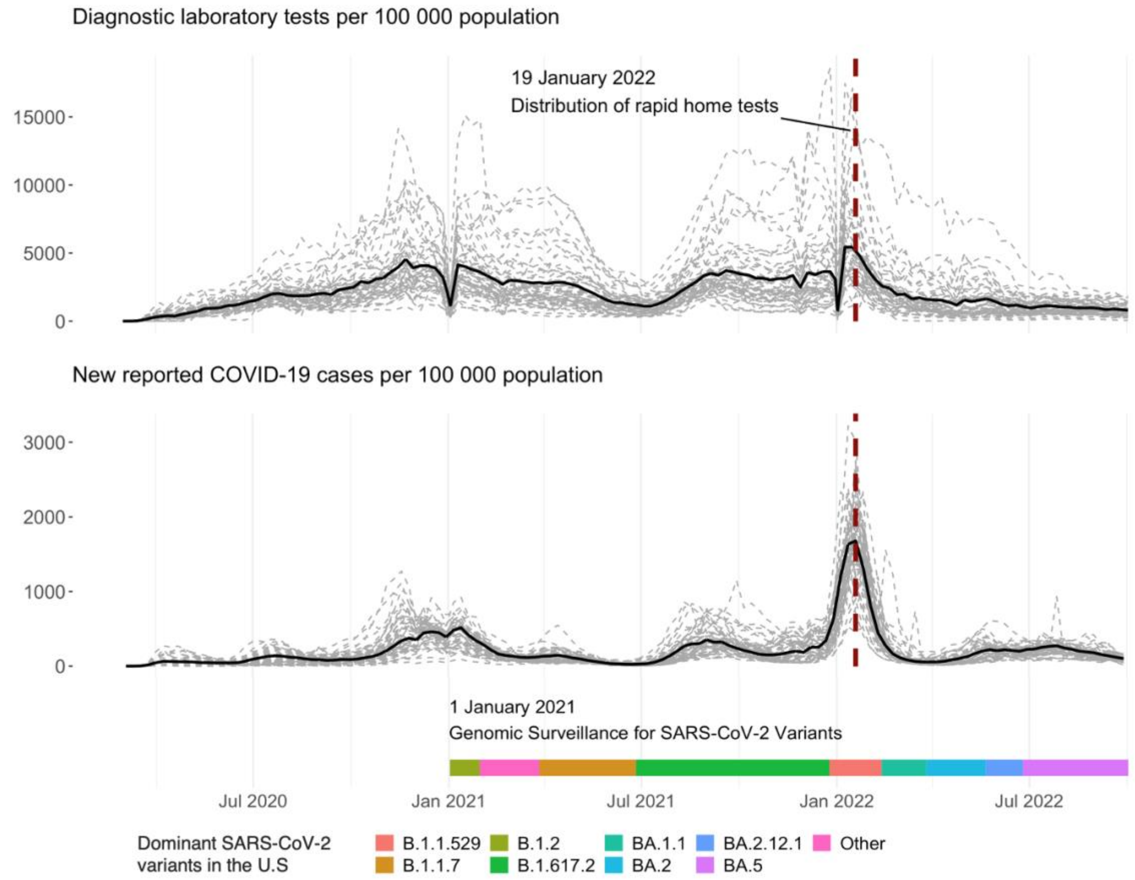


**Supplementary Figure 1. Diagnostic testing and reported new COVID-19 cases in the U.S between March 1 2020 and September 30 2022.**

The dashed gray lines represent weekly totals of diagnostic laboratory tests (top) and new reported COVID-19 cases (bottom) per 100,000 population by state. The solid black line shows total numbers for the United States. The colored horizontal bars represent time periods during which specific SARS-CoV-2 variants were dominant in the United States reported by the CDC as of January 1 2021. The dashed vertical line shows the date when distribution of rapid home tests was announced by the Biden administration.

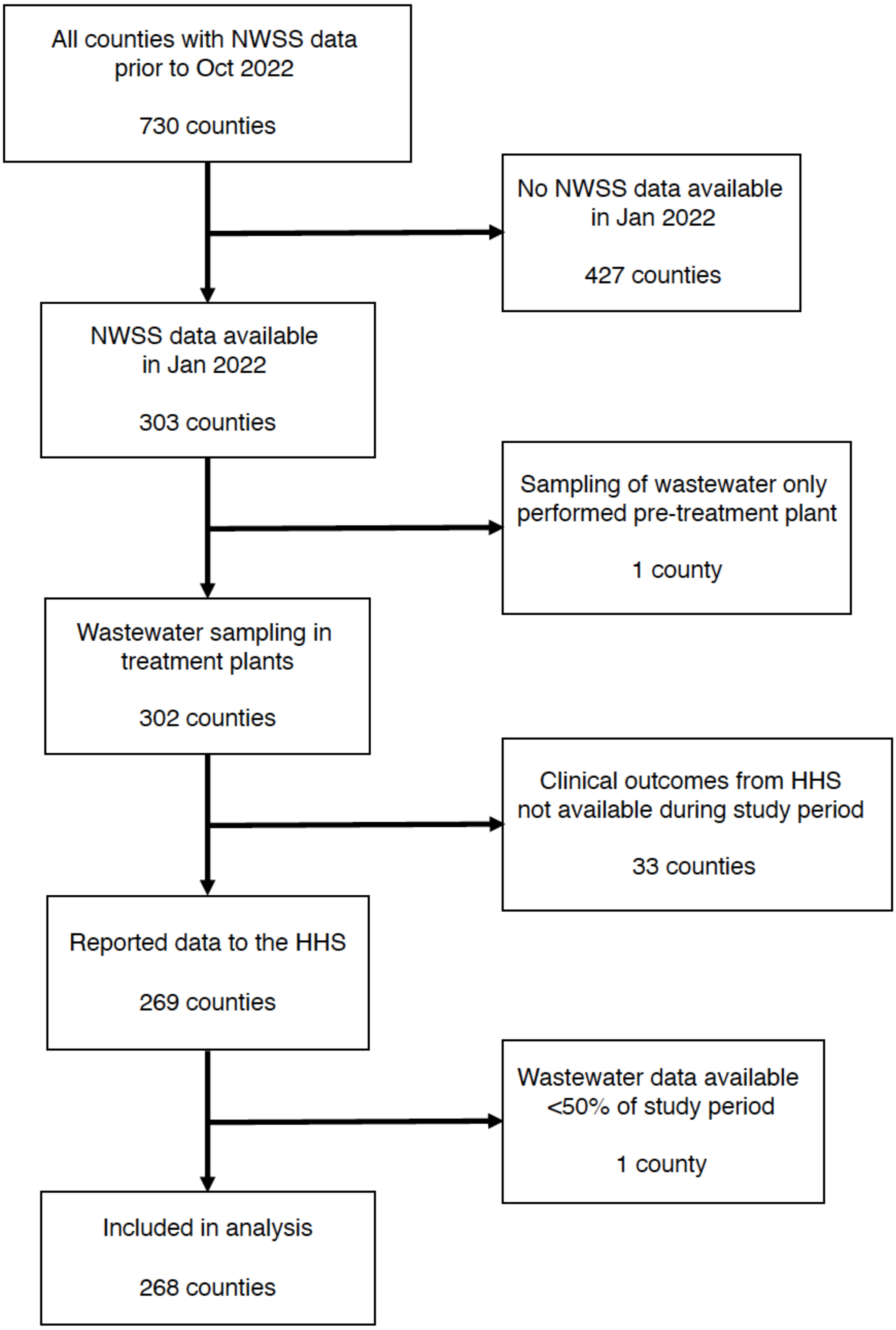


**Supplementary Figure 2. Selection of counties included in analysis.**

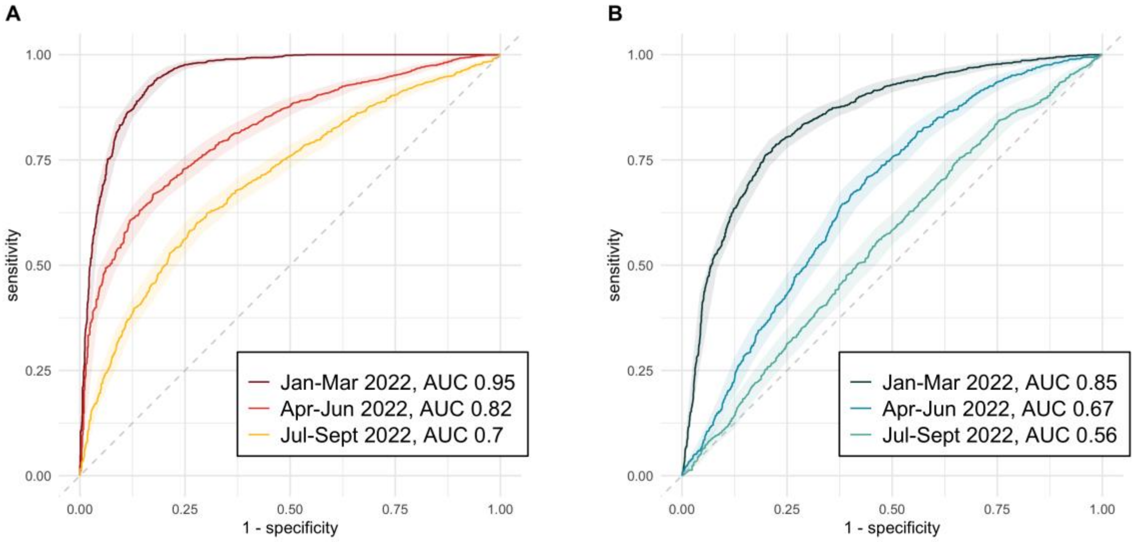


**Supplementary Figure 3. Performance of wastewater percentile in reference to clinical case metrics in small U.S. counties (n=230) stratified by calendar quartile of 2022.**

AUC of wastewater percentile in reference to (A) Current reported COVID-19 cases (≥200 per 100,000 population), (B) New hospital admissions in two weeks (≥10 per 100,000 population). Small counties were defined as counties with a total population less than 500,000 according to the 2021 U.S. Census.

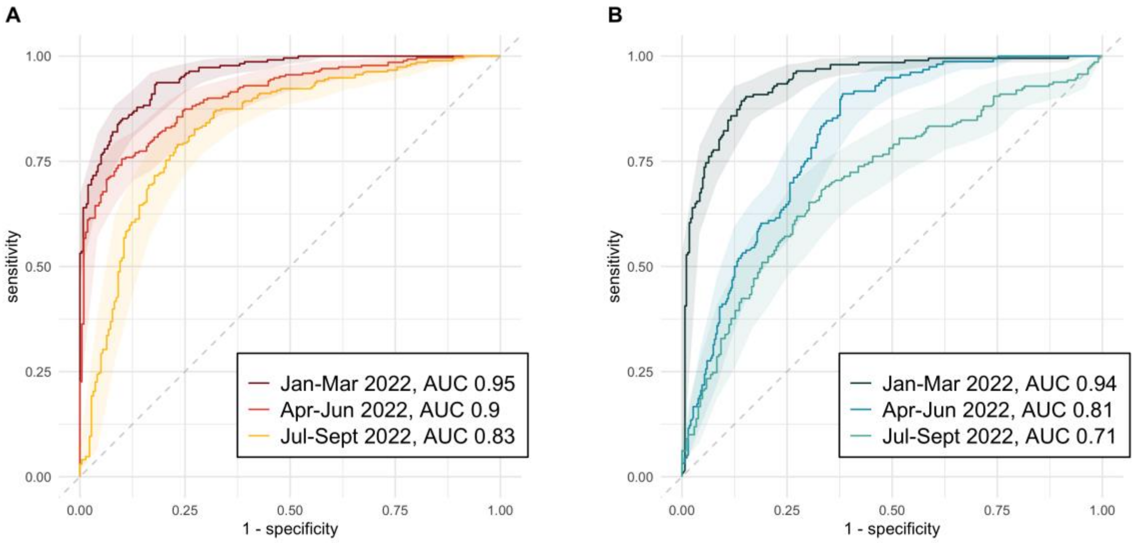


**Supplementary Figure 4. Performance of wastewater percentile in reference to clinical case metrics in large U.S. counties (n=38) stratified by calendar quartile of 2022.**

AUC of wastewater percentile in reference to (A) Current reported COVID-19 cases (≥200 per 100,000 population), (B) New hospital admissions in two weeks (≥10 per 100,000 population). Large counties were defined as counties with a total population equal to or greater than 500,000 according to the 2021 U.S. Census.

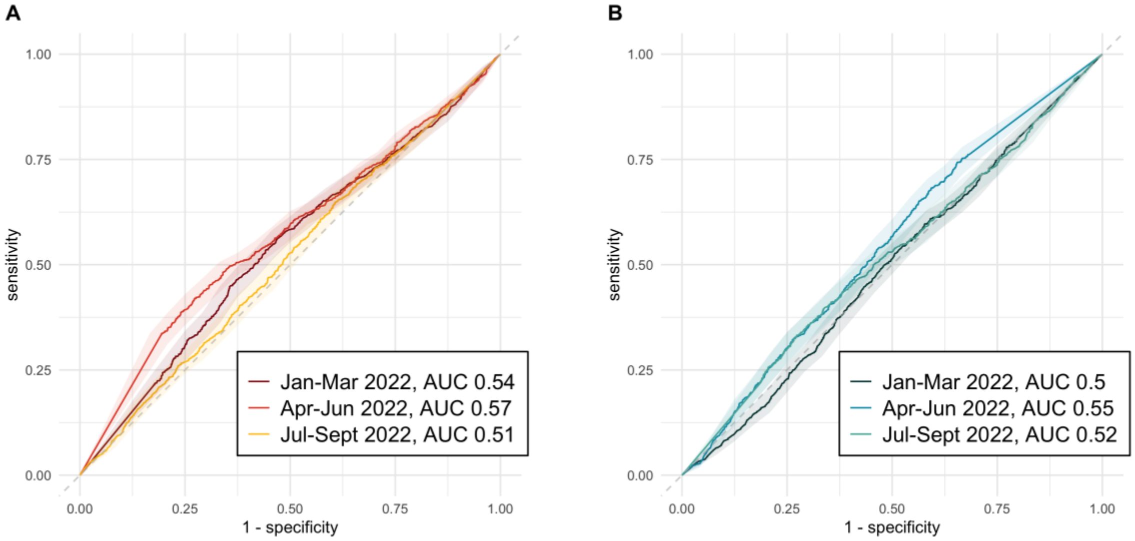


**Supplementary Figure 5. Performance of 15-day wastewater percent change in reference to clinical case metrics stratified by calendar quartile of 2022.**

AUC of 15-day wastewater percent change in reference to (A) Current reported COVID-19 cases (≥200 per 100,000 population), (B) New hospital admissions in two weeks (≥10 per 100,000 population).

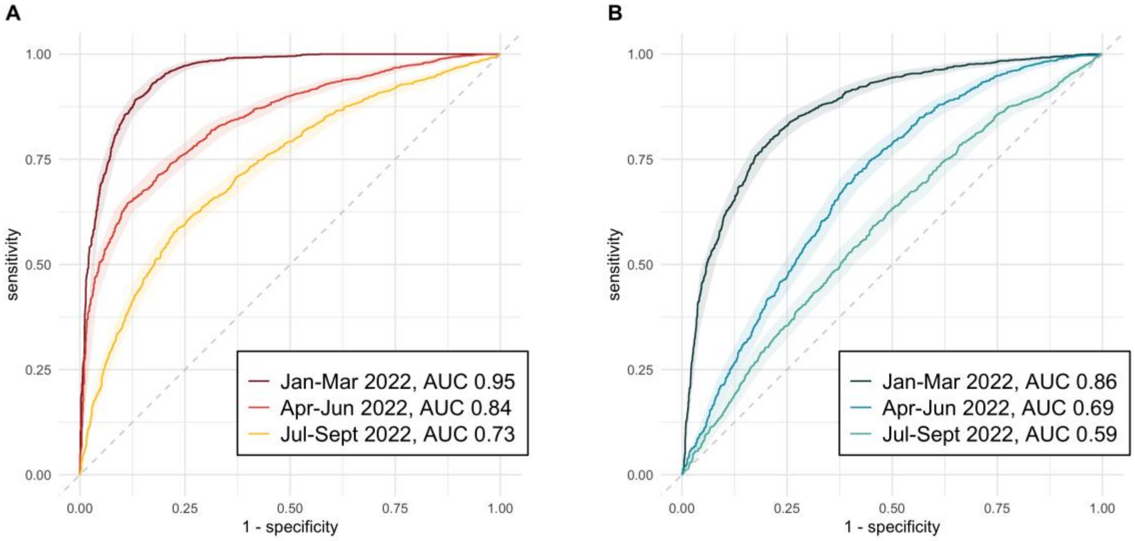


**Supplementary Figure 6. Performance of combined wastewater metrics in reference to clinical case metrics stratified by calendar quartile of 2022.**

AUC of combined wastewater metrics in reference to (A) Current reported COVID-19 cases (≥200 per 100,000 population), (B) New hospital admissions in two weeks (≥10 per 100,000 population). To estimate combined effects of wastewater metrics we used logistic regression accounting for wastewater percentile, percent change, and the interaction of the two.

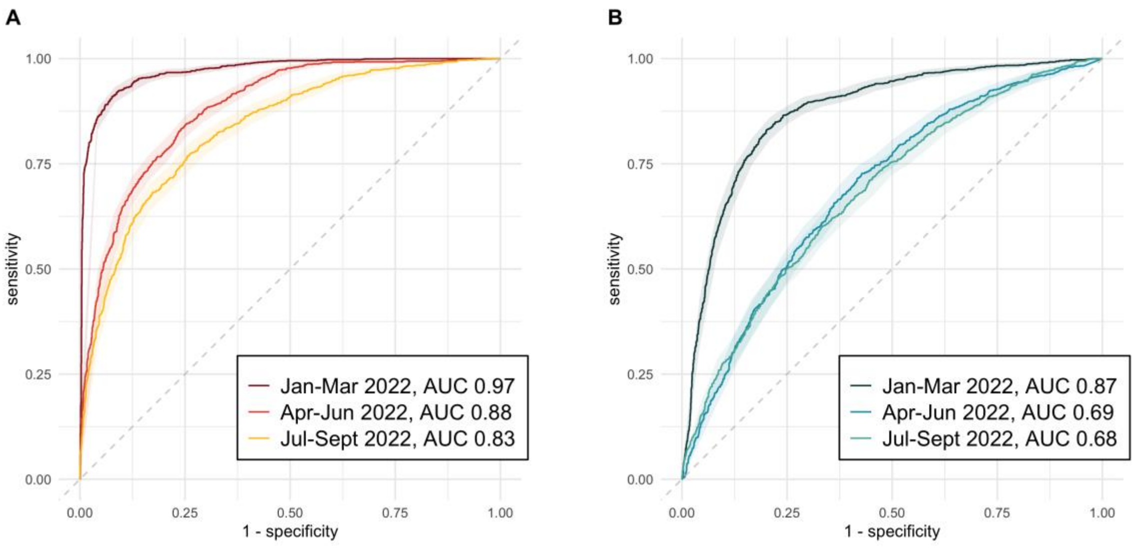


**Supplementary Figure 7. Performance of current reported COVID-19 case rates in reference to clinical case metrics stratified by calendar quartile of 2022.**

AUC of current reported case rates in reference to (A) reported COVID-19 cases in two weeks (≥200 per 100,000 population), (B) New hospital admissions in two weeks (≥10 per 100,000 population).

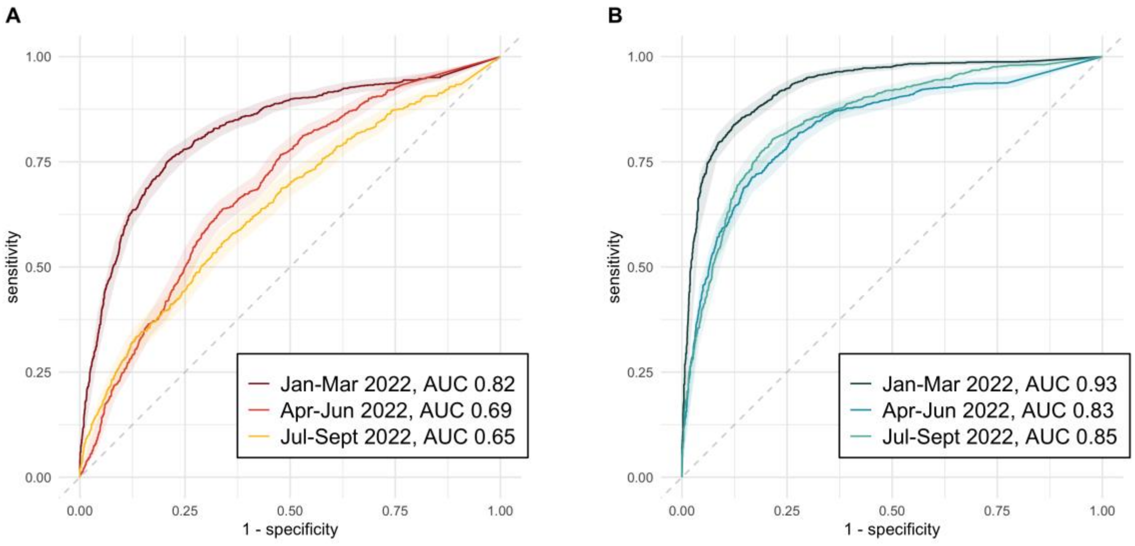


**Supplementary Figure 8. Performance of current COVID-19 hospital admission rate in reference to clinical case metrics stratified by calendar quartile of 2022.**

AUC of current hospitalization rates in reference to (A) reported COVID-19 cases in two weeks (≥200 per 100,000 population), (B) New hospital admissions in two weeks (≥10 per 100,000 population).
